# MUSCLE ENDURANCE, NEUROMUSCULAR FATIGABILITY AND COGNITIVE CONTROL DURING PROLONGED DUAL-TASK IN PEOPLE WITH CHRONIC OBSTRUCTIVE PULMONARY DISEASE: A CASE-CONTROL STUDY

**DOI:** 10.1101/2024.03.05.24303798

**Authors:** Cyril Chatain, Jean-Marc Vallier, Nicolas Paleiron, Fanny Cucchietti Waltz, Sofiane Ramdani, Mathieu Gruet

## Abstract

Recent studies suggest that, compared to healthy individuals, people with chronic obstructive pulmonary disease (pwCOPD) present a reduced capacity to perform cognitive-motor dual-task (CMDT). However, these studies were focused on short-duration CMDT offering limited insight to prolonged CMDT inducing fatigue, which can be encountered in daily life. The present study aimed to explore the effect of adding a cognitive task during repeated muscle contractions on muscle endurance, neuromuscular fatigability and cognitive control in pwCOPD compared to healthy participants Thirteen pwCOPD and thirteen age- and sex-matched healthy participants performed submaximal isometric contractions of the knee extensors until exhaustion in two experimental sessions: (1) without cognitive task and (2) with a concurrent working memory task (i.e., 1-back task). Neuromuscular fatigability (as well as central and peripheral components measured by peripheral magnetic stimulation), cognitive performance and perceived muscle fatigue were assessed throughout the fatiguing tasks. Independently to the experimental condition, pwCOPD exhibited lower muscle endurance compared to healthy participants (p=0.039), mainly explained by earlier peripheral fatigue and faster attainment of higher perceived muscle fatigue (p<0.05). However, neither effect of cognitive task (p=0.223) nor interaction effect (group × condition; p=0.136) was revealed for muscle endurance. Interestingly, cognitive control was significantly reduced only in pwCOPD at the end of CMDT (p<0.015), suggesting greater difficulty for patients with dual-tasking under fatigue. These findings provide novel insights into how and why fatigue develops in COPD in dual-task context, offering a rationale for including such tasks in rehabilitation programs.

## 1) INTRODUCTION

Chronic obstructive pulmonary disease (COPD) is a progressive disease characterized by airflow limitation associated with an abnormal inflammatory response of the lungs (Celli et al. 2004). In addition to its impact on lung function, COPD can also lead to systemic consequences, including neuromuscular impairments (Maltais et al. 2014) and cognitive deficits (Cleutjens et al. 2014).

Limb muscle dysfunction is well-documented in people with COPD (pwCOPD) and is considered as a key systemic consequence of the disease. Some direct or indirect neuromuscular adaptations resulting from the disease have been identified (e.g., muscle fiber shift and atrophy, reduced muscle oxidative capacity, mitochondrial dysfunction) leading to muscle weakness, increased fatigability and reduced muscle endurance which limit pwCOPD in daily life activities (Maltais et al. 2014; Gruet 2018). PwCOPD can also exhibit impairment in cognitive processing during dual tasking (Ozsoy et al. 2021), restraining them throughout everyday tasks.

Many daily activities involve the simultaneous accomplishment of cognitive and motor tasks, known as cognitive-motor dual-task situations (CMDT) such as talking while walking or identifying the front door key on a key ring while climbing the stairs. Although few studies have focused on the concept of dual tasks in pwCOPD (for review, see Rassam et al. 2023), they agree that patients have insufficient attentional resources to successfully perform both tasks simultaneously compared to healthy controls, exhibiting thus a higher dual-task interference. For instance, Morlino et al. (2017) showed that the time to perform Timed Up and Go test (a basic functional mobility test) increased to a greater extent from the single-task to the dual-task condition in pwCOPD compared to healthy controls. Similarly, Heraud et al. (2018) reported that stride time variability during a 15m walk was increased only in pwCOPD when transitioning from single to dual-task condition. These findings provide evidence that pwCOPD experience impairments in neural control during dual-tasking. Although the underlying mechanisms are not fully understood, some results suggest that the greater interference observed in patients may be explained by their inability to increase oxygenation in the prefrontal cortex (Hassan et al. 2020), a brain area both involved in cognitive tasks requiring attention, memory as well as motor planning and regulation (MacDonald et al. 2000; Tanaka and Watanabe 2012; Funahashi 2017).

While the aforementioned studies highlight the inability of pwCOPD to adequately perform short duration dual-tasks (i.e., from 10 to 30s duration), it is yet unknown how the disease affects their capacities to perform prolonged dual-tasks (i.e., involving a motor endurance task with concomitant cognitive task) which may be conducive to the development of both mental and neuromuscular fatigability.

Several studies have reported that imposing a cognitive task during a prolonged isometric contraction could lead to a decrease in muscle endurance (i.e., reduced time to task failure) both in young (Yoon et al. 2009; Mehta and Agnew 2012; Keller-Ross et al. 2014; Chatain et al. 2019) and old participants (Pereira et al. 2015; Shortz and Mehta 2017). For instance, Pereira et al. (2015) showed a shorter time to task failure during submaximal sustained elbow flexion in presence of high and low cognitive demand (i.e., subtraction by 13 and by 1, respectively) compared to a control condition without cognitive demand (i.e., 16 ± 8 min vs. 17 ± 4 min vs. 21 ± 7 min, respectively) in old women.

Other studies did not report any deleterious effect of the addition of cognitive demand during prolonged muscle contraction on muscle endurance (Mehta and Parasuraman 2014; Guzmán-González et al. 2020). Nevertheless, these studies showed that cognitive performance was significantly decreased (i.e., increased reaction time, decreased accuracy or both) during CMDT compared to the same cognitive task performed alone. Then, it appears that prolonged CMDT inevitably leads to a decrease in cognitive and/or motor performance compared to the equivalent motor or cognitive task performed alone.

Therefore, it seems plausible that people with impairment in neuromuscular function and cognitive processing such as pwCOPD (Dodd et al. 2010; Marillier et al. 2022) would experience even greater difficulties in performing prolonged CMDT. Prolonged CMDT are frequent in daily life activities and are also increasingly used for rehabilitation purposes to improve both motor and cognitive functions in a time-effective manner (Johansson et al. 2023; Rozenberg et al. 2023). A better understanding of the neuromuscular adaptations and cognitive control over the course of a prolonged fatiguing CMDT is thus essential before seeking to implement such interventions in the package of care of pwCOPD.

Accordingly, the aim of the current study was to assess the effects of adding cognitive demand (i.e., working memory task) to prolonged submaximal isometric contractions of knee extensors on muscle endurance performance (i.e., time to motor task failure) and cognitive control between pwCOPD and age- and sex-matched healthy participants. Our main hypothesis was that muscle endurance would be decreased for both groups in CMDT compared to control condition (i.e., motor task alone) and would be further decreased in pwCOPD compared to healthy controls. We also hypothesized that this larger decline in muscle endurance during CMDT in pwCOPD compared to healthy controls would be observed along with larger increase in neuromuscular fatigability and larger decrease in cognitive control.

## 2) METHODS

### 2.1) Participants

Nineteen pwCOPD and seventeen age- and sex-matched control participants were initially enrolled in this study. To ensure optimal pairing between groups, we first recruited pwCOPD and then control participants with the following rule: healthy volunteers were considered eligible if their age corresponded to that of a pwCOPD of the same sex within a ± 6-years range. However, six pwCOPD and four control participants were subsequently excluded from the data analysis for various reasons, such as fatiguing task not conducted to exhaustion, or the inability to find matched pwCOPD or control participants.

Main inclusion criteria were as follows : (1) forced expiratory volume in the first second (FEV_1_; evaluated by plethysmography) < 80% of predicted values (for pwCOPD only); (2) stable state (i.e., without exacerbations) for more than fifteen days (for pwCOPD only); (3) body mass index (BMI) < 30 kg.m^−2^; (4) mini-mental state examination (MMSE) score ≥ 26; (5) without known and uncontrolled chronic respiratory, cardiovascular, metabolic, renal or neuromuscular pathologies (for control participants only); (6) French participant able to provide written consent. Main non-inclusion criteria included : (1) pwCOPD treated with oral or systemic corticosteroids (> 0.5mg.kg^−1^.day^−1^ for more than seven days) (for pwCOPD only); (2) patient oxygen dependent (for pwCOPD only); (3) alcoholism (i.e., > 21 or 14 drinks/week for men and women, respectively); (4) psychiatric pathologies or history of behavioral disorders; (5) contraindication to peripheral nerve magnetic stimulation; (6) severe vision or hearing problems not corrected; (7) pregnant or lactating women; (8) regular vigorous physical activity with a frequency of more than three sessions per week.

This study received ethical approval from an ethic committee (CPP Ile de France III, 2019-A01986-51) and registered at www.clinicaltrials.gov (NCT04028973). All participants provided written informed consent prior to their participation in the study and were free to withdraw at any time. Each participant received financial compensation (150€) after completing the study.

### 2.2) Sample size calculation

At the moment of registering the clinical trial, there were only very few studies on the topic. We believed that the best way to estimate the effect of dual-task interference in COPD came from studies focusing on the effect of COPD on the performance of a psychomotor task requiring both cognitive and motor skills (i.e., Trail Making Test). Only one case-control study provided sufficient descriptive statistics to calculate an effect size (Dogra et al. 2014). Based on the results, the minimum required sample size was calculated as 64 participants (i.e., 32 in each group) for an effect size of 0.79, a probability level of 0.05, a statistical power level of 90% and assuming a 10% loss to follow up or study exit rate.

However, at the start of the inclusions, several results obtained from protocols closest to that of our study were published, allowing us to perform more relevant sample size calculation. This is notably the case of results from Ozsoy et al. (2021), reporting an index of cognitive dual-task interference during a muscle force production test between 35 pwCOPD and 27 healthy participants (dual-task interference = 63.2±26.2% for pwCOPD and 27.4±26.4% for healthy participants). Based on these results, the minimum required sample size was calculated as 24 participants (i.e., 12 in each group) for an effect size of 1.36 while keeping the same values for the other parameters (i.e., probability level, statistical power and study exit rate described in the aforementioned *a priori* analysis).

In summary, the poverty of literature on the fatigability of pwCOPD in dual-task conditions makes it difficult to accurately determine the required sample size to demonstrate a significant inter-group difference between conditions for our primary outcome of interest (i.e., time to task failure). It should be noted that the initial objective was to meet the first sample size calculation (i.e., 64 participants) but in light of logistical issues, mainly caused by the covid-19 pandemic, and the intermediate sample size calculation, we decided to stop the inclusions at 36 participants (19 pwCOPD and 17 healthy controls).

### 2.3) Experimental design

This study was carried out at two research institutions. The experiments involving pwCOPD were conducted at pulmonology department of Saint-Anne army hospital (Toulon, France) while the experiments involving control participants were carried out at the University of Toulon (La Garde, France). However, all experiments were performed exactly with the same materials and environmental conditions, and by the same experimenter (C.C.).

Each participant visited the respective laboratory on three separate occasions within a 2-week period. During the first visit, participants were familiarized with the study procedures and equipment and they were asked to complete several questionnaires, i.e.,: (1) Mini-Mental State Examination (MMSE) questionnaire (Folstein et al. 1975); (2) the Pittsburgh Sleep Quality Index (PSQI) questionnaire (Buysse et al. 1989); (3) the French version of the Baecke questionnaire (Vol et al. 2011); (4) the Manchester COPD-fatigue scale (MCFS) (Al-shair et al. 2009) and (5) the VQ11 questionnaire (Ninot et al. 2013). The two last questionnaires were completed only by pwCOPD.

MMSE questionnaire, which includes 30 items for determining the orientation, memory, language and visual-spatial capacities, is commonly used to assess the cognitive function. The maximum score is 30, with a score < 26 being considered to indicate mild cognitive impairments.

PSQI questionnaire was used to assess sleep quality. It comprises 19 self-reported questions providing a total score ranging from 0 (better) to 21 (worse). A total score ≥ 5 is associated with poor sleep quality.

French Baecke questionnaire was used to assess the level of habitual physical activity. It is composed of several questions about the frequency, duration and intensity of physical activity performed in three different components (i.e., sports practice, leisure time and daily life activities). For each component the score ranges from 1 to 5 and the sum of these components provides the total physical activity score ranging from 3 to 15 (a high score indicating a high level of physical activity).

MCFS is a valid and reliable scale used to quantify fatigue in pwCOPD. MCFS is a 27-item self-reported scale including three dimensions (i.e., physical, cognitive and psychosocial) to quantify the fatigue level. The total score ranges from 0 to 54 with a higher score associated with higher level of fatigue.

VQ11 is a valid French self-administered questionnaire used to assess the health-related quality of life specific to COPD and composed of 11 items distributed in three components (i.e., functional, psychological and relational). The total score ranges from 11 to 55 with a high score reflecting a poor quality of life.

During the two following experimental sessions, the muscle fatiguing tasks were performed, in a random order, with or without concomitant cognitive task (Figure 1a).

**Figure 1.**
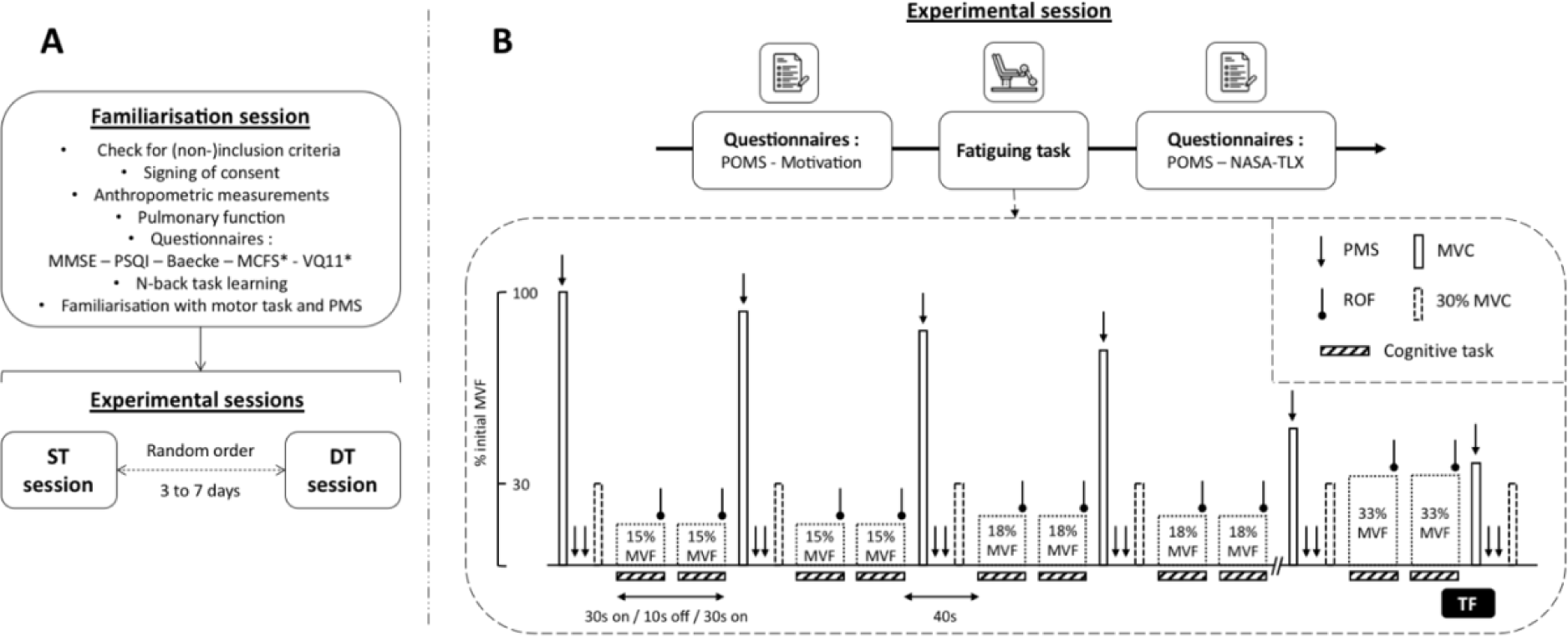
Overview of the experimental design (panel A), the experimental sessions and the fatiguing tasks (panel B). MSSE: Mini-mental state examination; PSQI: Pittsburgh sleep quality index; MCFS : Manchester COPD fatigue scale; VQ11 : Health-related quality of life; Baecke : Habitual physical activity; MVC : Maximal voluntary contraction; MVF : Maximal voluntary force; ROF : Rating-of-fatigue scale; PMS : Peripheral magnetic stimulation; TF : Task failure; ST : Single-task session; DT : Dual-task session; POMS : Profile of mood scale; NASA-TLX : NASA Task load index.

#### 2.3.1) Experimental sessions

The two experimental sessions were: (1) a single task (ST) session during which participants performed the muscle fatiguing task without concomitant cognitive task and (2) the dual-task (DT) session during which participants performed the same muscle fatiguing task with an added working memory task (i.e., 1-back task, see below). Each session was conducted on separate days, around the same time of day for a given participant and separated by at least 72h (maximum 7 days) to refrain from residual muscle fatigue effects. All participants were asked to refrain from caffeine and vigorous or prolonged physical activity 24h prior to each experimental session.

Upon arrival to the experimental center, participants completed the Profile of Mood States (POMS) (Terry et al. 2003) and motivation questionnaires (Matthews et al. 2001) (Figure 1b). For POMS questionnaire, participants were asked to rate “How are you feeling right now?” using 24 mood descriptors (e.g., depressed, sad, dynamic). For each descriptor participants had to answer using a 5-point Likert scale from 0 (not at all) to 4 (extremely). This questionnaire is divided into six subscales (i.e., fatigue, vigor, depression, confusion, tension and anger) each containing four mood descriptors. Motivation questionnaire is composed of two scales (i.e., success and intrinsic motivation) each containing seven descriptors (e.g., “I expect the exercise to be interesting”), for which the participants had to answer using a 5-point Likert scale from 0 (not at all) to 4 (extremely).

After questionnaires completion, if the experimental session was the DT session, participants accomplished the 1-back task during 60s. This “mental warm-up” was repeated until participants reach an overall accuracy ≥ 90%. Then, a standardized warm-up of the knee extensors (i.e., 4min30s intermittent isometric contractions at increasing force levels on the custom-built chair described below) was accomplished. After a 3-min recovery period, resting measurements of heart rate (HR) were obtained via a heart rate sensor (Polar H10, Polar Electro, Kempele, Finland).

Following HR recording, participants were asked to perform at least three maximal voluntary contractions (MVCs) of ∼5s duration interspaced by 1min of recovery. If a gradual increase was observed during the first three attempts, additional MVCs were performed to obtain a true maximal voluntary force (MVF). Strong verbal encouragements were given during each MVC to ensure maximal effort.

Then, a baseline assessment of neuromuscular function (see below) was performed. Finally, before beginning the muscle fatiguing task, participants were asked to rate their perceived muscle fatigue using the Rating-of-Fatigue (ROF) scale composed of 11 numerical points that range from 0 (not fatigued at all) to 10 (total fatigue and exhaustion) (Micklewright et al. 2017).

##### Muscle fatiguing task

Participants were set up in an adjustable custom-built chair with hips and knees at 130° (allowing optimal magnetic stimulation of the femoral nerve, e.g., Gruet et al. (2016), see below) and 90° of flexion, respectively. The lower right leg was attached to a force sensor (F2712 200 daN, Celians MEIRI, France) ∼5 cm above the malleoli of the ankle joint. The muscle fatiguing task consisted in sets of intermittent submaximal isometric contractions of the right knee extensors comprising two contractions of 30s, starting at 15% MVF for the first set and increasing by 3% MVF every two sets until task failure (Figure 2b). Task failure (TF) was defined as the inability to reach the target force level during four consecutives seconds. The force signal feedback was displayed on a TV screen placed in front of the participants using Acqknowledge software (version 4.1, Biopac Systems, Inc., Santa Barbara, CA, USA) and recorded at 2 kHz. The participants were asked to rate their perceived muscle fatigue using the ROF scale (Micklewright et al. 2017) throughout the fatiguing task, at the end of each 30-s contraction. Heart rate was continuously recorded during the fatiguing task.

**Figure 2.**
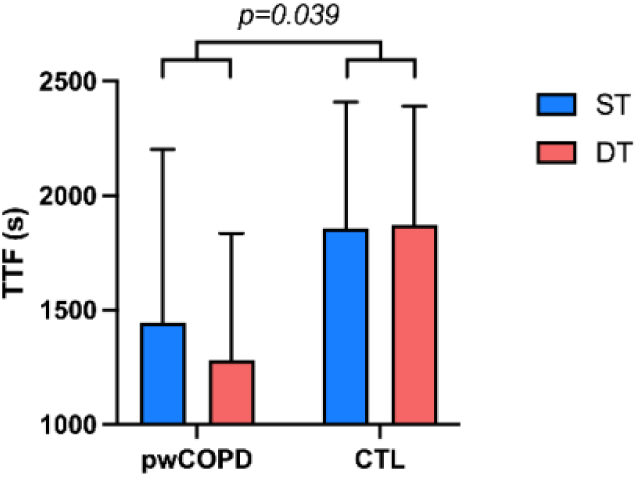
Effect of group and experimental condition on time to task failure (n=13). TTF : Time to task failure; pwCOPD : people with COPD; CTL : control participants. Data are presented as mean ± SD. The p value at the top of the figure corresponds to the main group effect.

##### Assessment of neuromuscular function

Assessments of neuromuscular function were performed at baseline, at the end of each sets and at TF (Figure 1b). The timing of the assessments was standardized and performed in 40s. Each neuromuscular assessment included a brief MVC of ∼5s duration. During each MVC one single superimposed stimulation was delivered to the right femoral nerve using a magnetic stimulator (Magstim200, The Magstim Company, Dyfed, UK) and a double-cone coil (50-mm outside diameter). The optimal stimulus site (i.e., position that induced the largest knee extensors twitch response) was obtained for each session and marked on the skin using a felt-tip pen to ensure reproducibility of stimulations throughout the experimental session. Following each MVC, two single stimulations were delivered on the relaxed muscle. The assessment of neuromuscular function ended by a submaximal contraction of ∼6s performed at 30% MVF with the aim to analyze the variability and temporal structure of force signals (Chatain et al. 2020, 2021) (data not shown as beyond the scope of this paper).

##### Cognitive task

During DT session, participants were asked to perform the muscle fatiguing task with a concomitant auditory 1-back task. We used 1-back task as stressing short-term memory is relevant to everyday life and also because we previously demonstrated (1) its feasibility in healthy subjects when used concomitantly to a motor task close to the one used in the present study (i.e., isometric quadriceps fatiguing task) and (2) that its addition to the motor task was able to induce a decline in motor performance (i.e., reduced muscle endurance), as compared to the realization of the motor task alone (Chatain et al. 2019). Participants were asked not to give a priority to motor or cognitive task and to perform both tasks as well as possible. The aim of this cognitive task was to evaluate whether a present stimulus is similar to the previous one. Stimuli were letters (i.e., J, L, M, Q, S, T, X, Z) presented using E-prime software (version 2.0, Psychology Software Tools, Sharsburg, MD, USA) and audio speakers. Participants’ responses were recorded using two computer mouses. If the current letter was similar to the previous one (i.e., match trial), the participants had to press the mouse computer held in the right hand. In reverse, if the current letter was different to the previous one (i.e., nonmatch trial), the participants had to press the mouse computer held in the left hand. Since the cognitive task was performed concomitantly to the motor task, the 1-back task was designed in such a way as to present stimuli only during the 30s-duration submaximal contractions. Each contraction comprised 14 letters presented every 2000ms and included 4 match trials. Participants were asked to provide their answers as accurately and quickly as possible. No feedback was given between each trial or at the end of a sequence. The sequences were different from one set to another for a given participant but were the same for a given set for all participants, ensuring optimal between-subjects comparisons.

### 2.4) Data analysis

#### 2.4.1) Neuromuscular data

Time to task failure (TTF) was calculated as the time between the beginning of the first submaximal contraction and TF. MVF was considered as the peak force reached during MVC. Potentiated peak twitch (Tw_p_; i.e., twitch obtained few seconds after the completion of MVC) amplitudes were obtained from evoked responses in the relaxed muscle. The two values of Tw_p_ were averaged for each neuromuscular function assessment. The voluntary level activation (VAL) was quantified by measurement of superimposed twitch amplitude (Tw_s_; i.e., twitch force added during the MVC by femoral nerve stimulation) and calculated using formula (1) including the correction proposed by Strojnik and Komi (1998) :

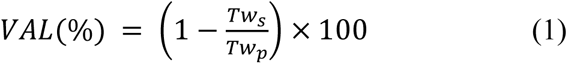

Force signals were filtered using a low-pass digital filter with cut-off frequency of 20 Hz before computation of MVF, Tw_p_ and Tw_s_.

#### 2.4.2) Cognitive performance

An indicator named D’, preferred to the percentage of good answer since it is not biased by individual response tendencies (Haatveit et al. 2010), was computed to quantify the cognitive performance using the following formula :

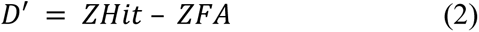

where Hit represents the proportion of correct matches (i.e., hits/(hits+misses)), FA (false alarms) represents the proportion of match responses on nonmatch trials (i.e., false alarms/(false alarms + correct negative)) and Z represents the inverse of the standard normal distribution. In case of perfect scores, Hit and FA were adjusted using the formula (3) and (4), respectively :

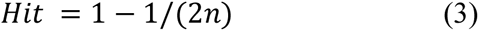

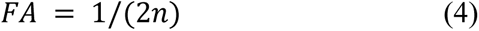

where n correspond to the number of total hits or false alarms. Accordingly the highest possible D’ value was +3.49 whereas the lowest value was −3.49.

The mean reaction time and the number of non-answers were also computed for each set. Responses given with a reaction time below 100ms, corresponding to an anticipation, were excluded from computation and were considered as non-answer.

All indicators (i.e., D’, reaction time, non-answer) were computed from two sequences of 14 letters presented during one set (i.e., from a sequence of 28 letters) in order to have a sufficient number of items to obtain a robust indicator of cognitive performance.

#### 2.4.3) Heart rate and perceived fatigue

For each 30s-duration submaximal contraction HR was averaged across 10s window centered in the middle of the contraction (i.e., excluding 10 first and last seconds). Then, two HR values obtained for both contractions composing a set were averaged in order to obtain one HR value for each set.

Similarly, ROF scores obtained at the end of each submaximal contraction were averaged to capture one score of perceived fatigue for each set.

#### 2.4.4) Isotime comparisons

Since muscle fatiguing tasks were performed until task failure, and consequently had different durations between participants and conditions, our results were compared using isotime comparisons (Nicolò et al. 2019). These analyses were necessary to interpret the dynamics of different parameters at the exact same amount of work between participants and conditions.

For this purpose, for each couple of participants (i.e., one COPD patient and one sex and age-matched control participant), we selected the shortest of four fatiguing tasks as reference task allowing us to determine the different time points : (1) Pre or 0% which corresponds to measures performed before or during the first set of the fatiguing task; (2) 25% which corresponds to measures performed at quarter of the fatiguing task duration; (3) 50% which corresponds to measures performed at half of the fatiguing task duration; (4) 75% which corresponds to measures performed at three-quarter of the fatiguing task duration; (5) 100% or TF which corresponds to measures performed at TF or during the last set of the fatiguing task entirely completed. Finally, we used exactly the same sets or neuromuscular assessments for the three other fatiguing tasks. In other words, results for Pre (or 0%), 25%, 50%, 75% and 100% time points for all parameters were compared at the same exercise time between conditions and between a COPD patient and his sex age-matched control participant. Note that 100% and TF time points are only merged for the shortest of the four fatiguing tasks (i.e., for the three other tasks TF and 100% correspond to different time points) and consequently only comparisons for TF time point were different in terms of exercise time.

If no neuromuscular assessment or set corresponded to exactly 25%, 50% or 75% of the duration of the shortest fatiguing task, the nearest was considered.

#### 2.4.5) Statistical analyses

All statistical procedures were performed on SPSS (version 27.0, IBM Corp., Armonk, NY, USA). For all variables, normality was inspected using Shapiro-Wilk test. Participants’ characteristics were compared between groups using independent sample t-test or Mann-Whitney test when conditions for the application of parametric test were not met. To detect effects of group, condition or interaction (group × condition), TTF and scores from motivation and NASA-TLX questionnaires were compared using linear mixed models (LMMs) with one within-subject factor (condition) and one between-subject factor (group). The same statistical procedure, with one within-subject factor (time) and one between-subject factor (group), was used for indicators of cognitive control (i.e., D’, reaction time, non-answer) to detect effects of group, time or interaction (time × group). To detect effects of time, group, condition or any interactions for all other variables recorded during fatiguing tasks (i.e., neuromuscular parameters, HR and ROF scores) and scores from the POMS questionnaire, LMMs with two within-subject factors (time and condition) and one between-subject factor (group) were used. The fixed factors of the LMMs were time, group, condition and all interactions between these factors. For all LMMs, a random effect structured by subjects was included. If a main effect or interaction was detected, LMMs were followed by pairwise comparison tests using the Bonferroni correction. LMMs were used because they allow to analyze repeated measures data with missing values and are robust to violations of distributional assumptions (Schielzeth et al. 2020). LMMs also provide greater statistical power that traditional analyses of variance (Ma et al. 2012). All data are presented as mean ± SD within the text and figures. Mean differences (MD) and 95% confidence intervals (95%CI) were also computed. For all analyses, statistical inferences were drawn at 0.05 level of significance.

## 3) RESULTS

### 3.1) Subjects characteristics

Characteristics of the 26 participants (14 men and 12 women) retained for analyses are given in Table 1. No differences between groups were reported for age and anthropometric measures. PwCOPD had lower resting pulmonary function outcomes and resting pulse oxygen saturation than healthy controls while resting HR was not different (Table 1). Regarding the neuromuscular parameters measured at rest, MVF, VAL and Tw_p_ were not different between groups. MMSE scores were not different between groups, while scores for Baecke (lower in pwCOPD) and PSQI (higher in pwCOPD) questionnaires were significantly different. According to the PSQI cut-off score, 38% and 77% of healthy participants and pwCOPD had poor sleep quality, respectively.

**Table 1.** Characteristics of participants retained for analyses. pwCOPD: People with COPD; CTL: Control participants; BMI: Body mass index; FEV_1_: Forced expiratory volume in 1 second; FVC: Forced vital capacity; RV : Residual volume; TLC: Total lung capacity; SpO_2_: arterial O_2_ saturation; HR: Heart rate; MVF: Maximal voluntary force; VAL: Voluntary activation level; Tw_p_: Potentiated peak twitch; MMSE: Mini-mental state examination; PSQI: Pittsburgh sleep quality index; MCFS : Manchester COPD fatigue scale; VQ11 : Health-related quality of life; Baecke : Habitual physical activity. For questionnaires, data are presented as median ± interquartile range. For parametric and non-parametric comparisons, the effect size are expressed as Cohen’s D or Rosenthal’s r, respectively. *(n=12) **(n=10).

| Variables | pwCOPD (n=13) |  | CTL (n=13) |  | p value |  | effect size |  |
| --- | --- | --- | --- | --- | --- | --- | --- | --- |
|  | Absolute | % Predicted | Absolute | % Predicted | Absolute | % Predicted | Absolute | % Predicted |
| Age (years) | 68.6 ± 7.4 | / | 67.4 ± 6.6 | / | 0.659 | / | 0.176 | / |
| Height (cm) | 165.9 ± 8.4 | / | 171.8 ± 8.2 | / | 0.085 | / | 0.704 | / |
| Weight (kg) | 68.8 ± 10.9 | / | 73.9 ± 16.5 | / | 0.356 | / | 0.370 | / |
| BMI (kg.m<sup>-2</sup>) | 25.0 ± 3.8 | / | 24.9 ± 4.6 | / | 0.941 | / | 0.029 | / |
| FEV<sub>1</sub> (L) | **1.5 ± 0.5** | **57.6 ± 12.1** | **3.3 ± 0.8** | **121.2 ± 14.8** | **<0.001** | **<0.001** | 2.651 | 4.701 |
| FVC (L) | **2.5 ± 0.9** | **73.4 ± 14.9** | **4.2 ± 1.0** | **125.8 ± 17.9** | **<0.001** | **<0.001** | 1.851 | 3.186 |
| FEV<sub>1</sub>/FVC (%) | **62.0 ± 15.0** | **79.9 ± 18.8** | **77.5 ± 3.9** | **100.3 ± 5.5** | **0.002** | **0.001** | 1.404 | 1.466 |
| RV (L) | 3.9 ± 2.8 | 196.5 ± 137.9* | / | / | / | / | / | / |
| TLC (L) | 6.7 ± 3.0 | 119.8 ± 52.0* | / | / | / | / | / | / |
| SpO<sub>2</sub> (%) | **96.6 ± 1.0** | / | **97.6 ± 1.2** | / | **0.027** | / | 0.923 | / |
| HR (bpm) | 82.3 ± 16.3 | / | 73.8 ± 8.3 | / | 0.108 | / | 0.654 | / |
| MVF (N.m) | 138.5 ± 54.8 | / | 177.2 ± 59.7 | / | 0.098 | / | 0.675 | / |
| VAL (%) | 93.8 ± 3.5** | / | 91.5 ± 10.4** | / | 0.971 | / | 0.170 | / |
| Tw<sub>p</sub> (N.m) | 38.0 ± 12.6** | / | 43.3 ± 15.4** | / | 0.404 | / | 0.382 | / |
| MMSE (0-30) | 29.0 ± 1.0 | / | 28.0 ± 2.0 | / | 0.687 | / | 0.083 | / |
| Baecke (3-15) | **6.9 ± 1.5** | / | **8.6 ± 1.4** | / | **0.014** | / | 1.044 | / |
| PSQI (0-21) | **7.0 ± 7.0** | / | **4.0 ± 4.0** | / | **0.002** | / | 0.586 | / |
| VQ11 (11-55) | 27.0 ± 13.0 | / | / | / | / | / | / | / |
| MCFS (0-54) | 24.5 ± 19.5 | / | / | / | / | / | / | / |

### 3.2) Time to task failure

Significant main group effect [MD=501.9; 95%CI=[28.2;975.6]; F(1,24)=4.78; *p*=0.039] was found for TTF without main condition effect [MD=72.4; 95%CI=[−47.0;191.7]; F(1,24)=1.57; *p*=0.223] or interaction effect [F(1,24)=2.39; *p*=0.136] (pwCOPD ST : 1444±759; pwCOPD DT : 1282±554; CTL ST : 1856±553; CTL DT 1873±518; Figure 2).

### 3.3) Neuromuscular fatigability

MVF decreased significantly during the fatiguing tasks [(F(5,192.9)=74.38; *p*<0.001] without main effect of group [MD=37.1; 95%CI=[−3.7;77.9]; F(1,24.0)=3.52; *p*=0.073], condition [MD=0.7; 95%CI=[−5.6;4.1]; F(1,42.7)=0.09; *p*=0.764] or any interaction (Figure 3a). Significant main effect of time was found for Tw_p_ [F(5,145.5)=61.4; *p*<0.001] without main effect of group [MD=6.5; 95%CI=[−2.8;15.8]; F(1,18.3)=2.15; *p*=0.159] or condition [MD=0.3; 95%CI=[−2.9;2.3]; F(1,27.7)=0.056; *p*=0.815]. Significant effect was also revealed for (group × time) interaction [F(5,145.5)=3.57; *p*=0.004] without any other interaction. Tw_p_ was significantly lower for pwCOPD at 100% time point (MD=10.2; 95%CI=[0.5;19.9]; *p_Bonferroni_*=0.04) (Figure 3b). VAL decreased significantly during the fatiguing tasks [F(5,136.3)=6.08; *p*<0.001] with main condition effect [MD=2.4; 95%CI=[0.15;4.6]; F(1,34.3)=4.69; *p*=0.037] and without main effect of group [MD=0.7; 95%CI=[−6.9;8.3]; F(1,18.0)=0.04; *p*=0.844] or any interaction (Figure 3c).

**Figure 3.**
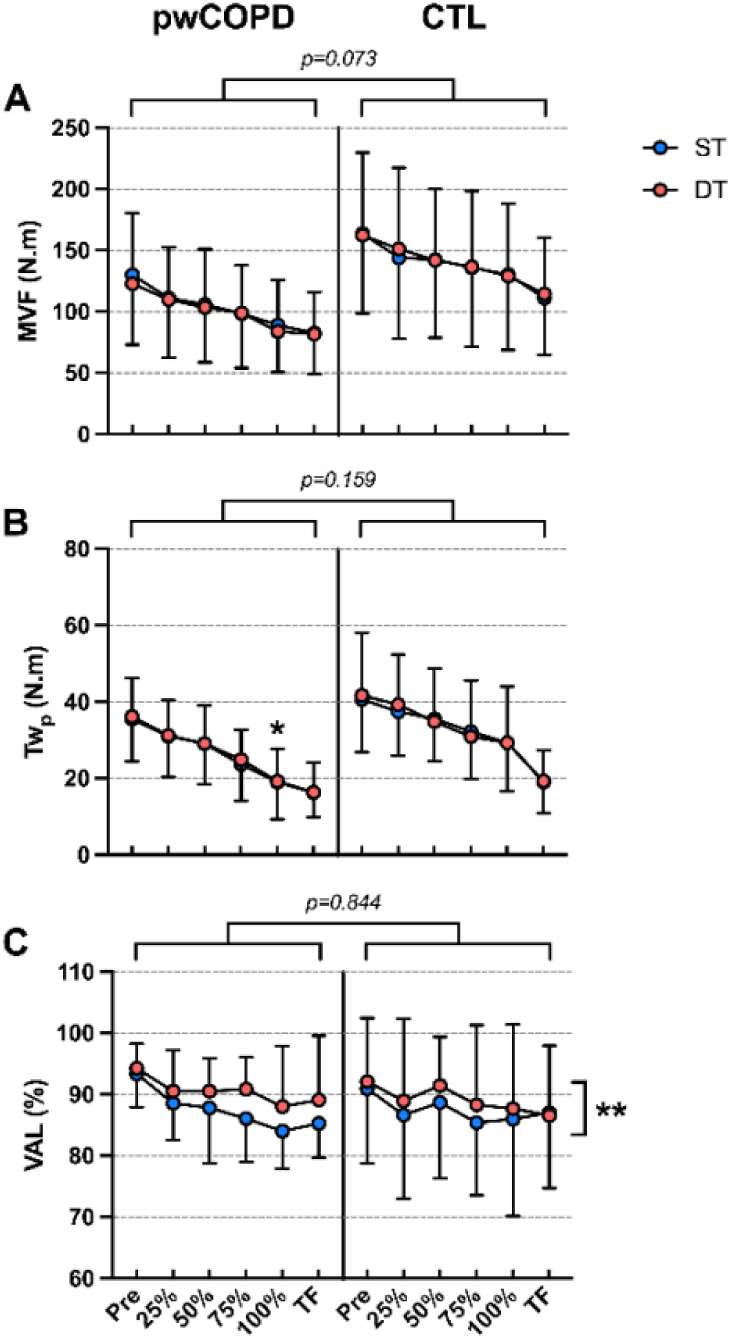
Maximal voluntary force (MVF, panel A), potentiated peak twitch (Twp, panel B) and voluntary activation level (VAL, panel C) of the knee extensors during the fatiguing tasks in people with COPD (pwCOPD) and control (CTL) participants in single task (ST) and dual-task (DT) conditions. Pre : before the fatiguing task; 25% : at 25% of the duration of shortest test; 50% : at 50% of the duration of shortest test; 75% : at 75% of the duration of shortest test; 100% : at 100% of the duration of shortest test and TF : at task failure. Data are presented as mean ± SD (n=10 except for MVF where n=13). The p values at the top of the panels correspond to the main group effect. *: significantly lower compared to 100% time point in CTL; **: significant effect of experimental condition.

### 3.4) Perceived muscle fatigue

ROF score increased significantly throughout the fatiguing tasks [F(6,216.6)=162.7; *p*<0.001] without main effect of group [MD=0.8; 95%CI=[−0.3;2.0]; F(1,22.8)=2.32; *p*=0.142] or condition [MD=0.04; 95%CI=[−0.57;0.49]; F(1,40.9)=0.02; *p*=0.883]. Significant effect was revealed for (group × time) interaction [F(6,216.6)=5.61; *p*<0.001] without any other interaction. ROF score was significantly higher for pwCOPD at 100% time point (MD=1.6; 95%CI=[0.3;2.9]; *p_Bonferroni_*=0.017) (Figure 4).

**Figure 4.**
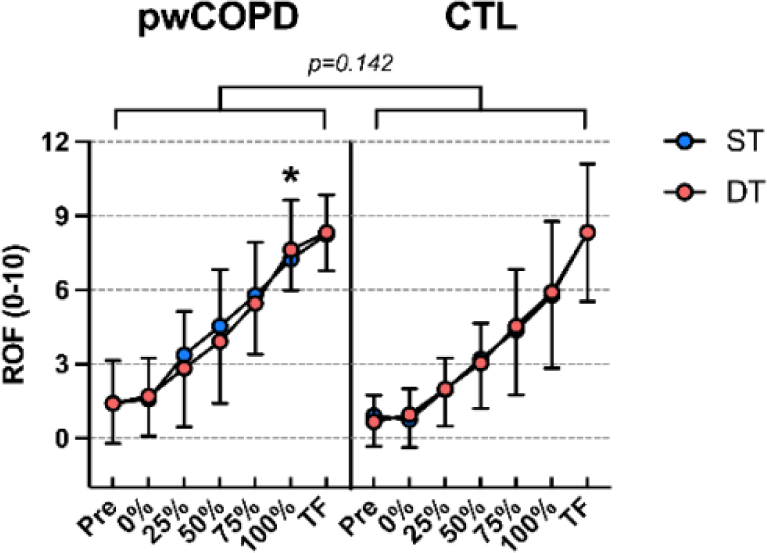
Rating of fatigue (ROF) scores during the fatiguing task in people with COPD (pwCOPD) and control (CTL) participants in single task (ST) and dual-task (DT) conditions. Pre : before the fatiguing task; 0% : at the end of the first set of fatiguing task; 25% : at the end of set corresponding to 25% of the duration of shortest test; 50% : at the end of set corresponding to 50% of the duration of shortest test; 75% : at the end of set corresponding to 75% of the duration of shortest test; 100% : at the end of set corresponding to 100% of the duration of shortest test and TF : at task failure. Data are presented as mean ± SD (n=12). The p value at the top of the figure corresponds to the main group effect. *: significantly higher compared to 100% time point in CTL.

### 3.5) Cognitive performance

D’ decreased significantly throughout the fatiguing tasks [F(5,69.5)=3.01; *p*=0.016] with main group effect [MD=0.47; 95%CI=[0.06;0.89]; F(1,24.0)=5.46; *p*=0.028] and interaction effect [F(5,69.5)=2.87; *p*=0.021]. D’ was significantly lower for pwCOPD at 75% (MD=-0.72; 95%CI=[−1.26;-0.17]; *p_Bonferroni_*=0.011), 100% (MD=-0.77; 95%CI=[−1.31;-0.22]; *p_Bonferroni_*=0.006) and TF (MD=-0.86; 95%CI=[−1.40;-0.31]; *p_Bonferroni_*=0.002) time points (Figure 5a). LMM revealed no significant changes of reaction time throughout the fatiguing tasks [F(5,69.4)=0.836; *p*=0.529], with no significant effect of group [MD=15.2; 95%CI=[−79.1;48.8]; F(1,24.8)=0.239; *p*=0.629] or interaction [F(5,69.4)=0.29; *p*=0.917] (Figure 5b). No significant main time effect [F(5,70.7)=1.86; *p*=0.113], main group effect [MD=1.74; 95%CI=[−0.38;3.87]; F(1,24.1)=2.87; *p*=0.103] or interaction effect [F(5,70.7)=1.45; *p*=0.217] was found for the number of non-answers (Figure 5c).

**Figure 5.**
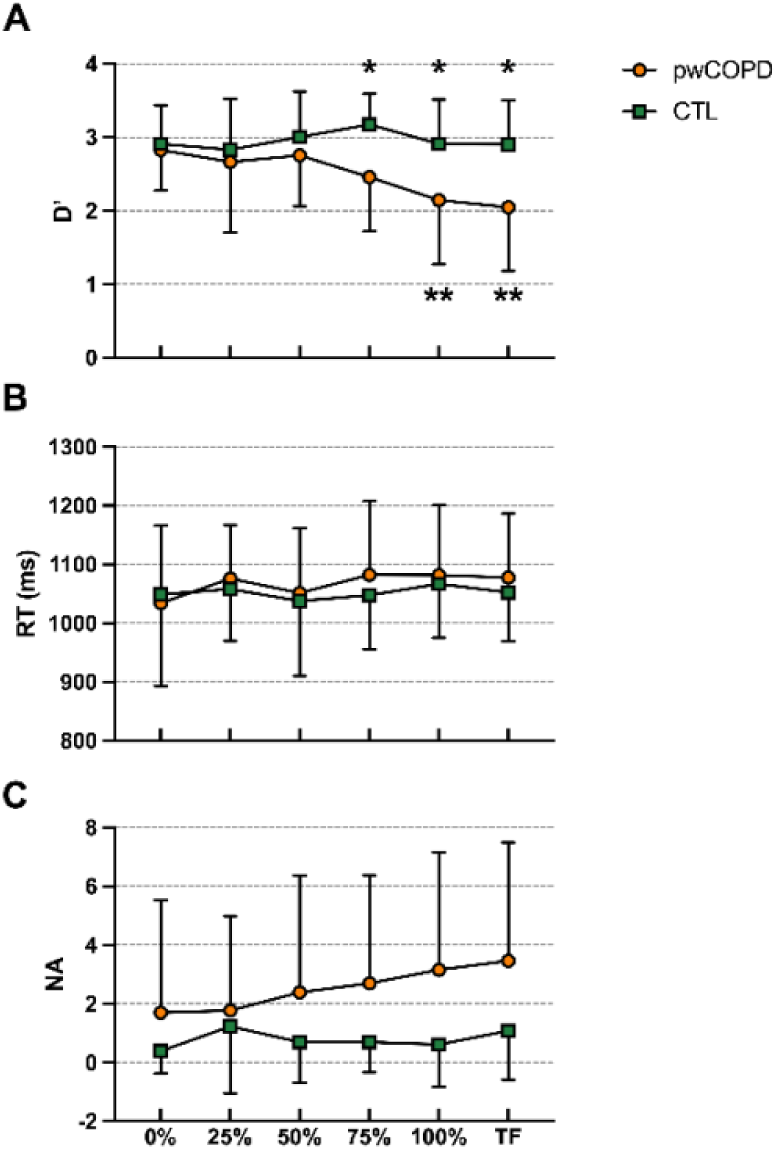
D’ (panel 1), reaction time (RT, panel B) and number of non-answers (NA, panel C) for 1-back task performed during the dual-task condition for people with COPD (pwCOPD) and control (CTL) participants. 0% : during the first set of the fatiguing task; 25% : at set corresponding to 25% of the duration of shortest test; 50% : at set corresponding to 50% of the duration of shortest test; 75% : at set corresponding to 75% of the duration of shortest test; 100% : at set corresponding to 100% of the duration of shortest test and TF : at the last set entirely completed. Data are presented as mean ± SD (n=13).*: significant difference between pwCOPD and CTL; **: significant difference with 0% time point in pwCOPD.

### 3.6) Heart rate

HR increased significantly throughout the fatiguing tasks [F(6,208.9)=15.3; *p*<0.001] with main condition effect [MD=3.1; 95%CI=[0.9;5.3]; F(1,44.0)=8.01; *p*=0.007] and without main effect of group [MD=6.2; 95%CI=[−4.8;17.2]; F(1,22.06)=1.37; *p*=0.255] or any interaction (Figure 6).

**Figure 6.**
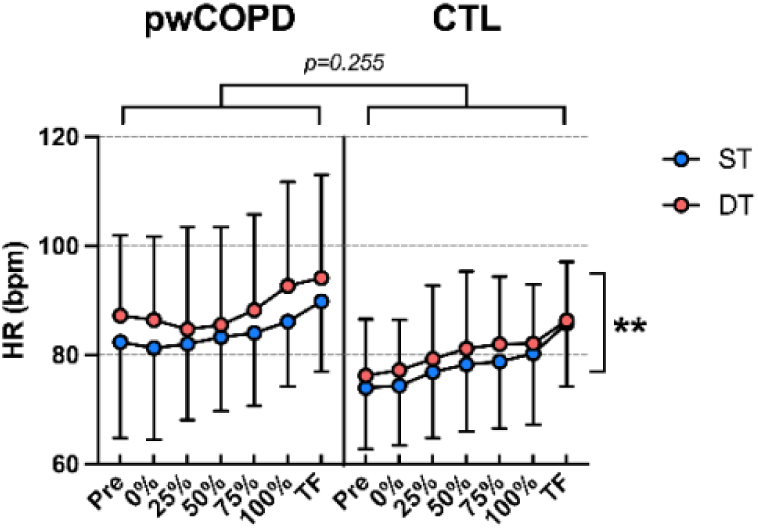
Heart rate (HR) during the fatiguing task for people with COPD (pwCOPD) and control (CTL) participants during single task (ST) and dual task (DT) conditions. Pre : before the fatiguing task; 0% : during the first set of fatiguing task; 25% : during the set corresponding to 25% of the duration of shortest test; 50% : during the set corresponding to 50% of the duration of shortest test; 75% : during the set corresponding to 75% of the duration of shortest test; 100% : during set corresponding to 100% of the duration of shortest test and TF : during the last set entirely completed. Data are presented as mean ± SD (n=12). The p value at the top of the figure corresponds to the main group effect. **: significant effect of experimental condition.

### 3.7) Questionnaires

#### 3.7.1) Motivation

Intrinsic motivation was not significantly different between groups [MD=0.73; 95%CI=[−1.6;3.1]; F(1,24)=0.41; *p*=0.528], conditions [MD=0.58; 95%CI=[−0.4;1.5]; F(1,24)=1.55; *p*=0.225] with the absence of interaction effect [F(1,24)=1.17; *p*=0.291] (pwCOPD ST : 21.2±2.3; pwCOPD DT : 20.1±3.9; CTL ST : 21.4±3.0; CTL DT 21.3±3.2). In the same way, success motivation was not different between groups [MD=0.15; 95%CI=[−3.5;3.2]; F(1,24)=0.01; *p*=0.924], conditions [MD=0.39; 95%CI=[−1.0;1.8]; F(1,24)=0.32; *p*=0.579] and without effect of interaction [F(1,24)=0.01; *p*=0.911] (pwCOPD ST : 15.6±4.4; pwCOPD DT : 15.3±5.7; CTL ST : 15.5±3.2; CTL DT 15.1±4.1).

#### 3.7.2) Mood

All detailed results for POMS questionnaire are available in Table 2. In short, three-way mixed model revealed a significant main group effect for anger [MD=0.8; 95%CI=[0.04;1.58]; F(1,24.0)=4.7; *p*=0.04], fatigue [MD=1.6; 95%CI=[0.1;3.2]; F(1,24.6)=4.63; *p*=0.041] and tension [MD=1.1; 95%CI=[0.07;2.2]; F(1,24.0)=4.8; *p*=0.038] subscales, with scores systematically higher for pwCOPD.

**Table 2.** Scores of Profil of Mood States questionnaire (POMS) before (Pre) and after (Post) single (ST) and dual (DT) fatiguing tasks in people with COPD (pwCOPD) and control participants (CTL). # : significant (time x group) interaction effect; ## : significant (condition x group) interaction effect. Data are presented as median ± interquartile range.

|  |  | pwCOPD |  | CTL |  |  |  |
| --- | --- | --- | --- | --- | --- | --- | --- |
|  |  | ST | DT | ST | DT | p value |  |
| Anger | Pre | 0.0 ± 0.0 | 0.0 ± 3.0 | 0.0 ± 0.0 | 0.0 ± 0.0 | Time | 0.069 |
|  |  |  |  |  |  | Group | **0.040** |
|  | Post | 0.0 ± 0.0 | 0.0 ± 0.0 | 0.0 ± 0.0 | 0.0 ± 0.0 | Condition | 0.101 |
|  |  |  |  |  |  | Interaction | none |
| Confusion | Pre | 0.0 ± 2.0 | 2.0 ± 4.0 | 0.0 ± 0.0 | 0.0 ± 2.0 | Time | 0.318 |
|  |  |  |  |  |  | Group | 0.084 |
|  | Post | 0.0 ± 1.0 | 1.0 ± 3.0 | 0.0 ± 0.0 | 0.0 ± 2.0 | Condition | **0.002** |
|  |  |  |  |  |  | Interaction | none |
| Depression | Pre | 0.0 ± 1.0 | 1.0 ± 2.0 | 0.0 ± 0.0 | 0.0 ± 0.0 | Time | **0.010** |
|  |  |  |  |  |  | Group | 0.070 |
|  | Post | 0.0 ± 0.0 | 0.0 ± 0.0 | 0.0 ± 0.0 | 0.0 ± 0.0 | Condition | 0.060 |
|  |  |  |  |  |  | Interaction | **# - ##** |
| Fatigue | Pre | 1.0 ± 4.0 | 3.0 ± 4.0 | 0.0 ± 1.0 | 0.0 ± 1.0 | Time | **<0.001** |
|  |  |  |  |  |  | Group | **0.041** |
|  | Post | 3.0 ± 3.0 | 5.0 ± 4.0 | 2.0 ± 4.0 | 2.0 ± 3.0 | Condition | 0.164 |
|  |  |  |  |  |  | Interaction | none |
| Tension | Pre | 0.0 ± 2.0 | 2.0 ± 5.0 | 0.0 ± 1.0 | 0.0 ± 1.0 | Time | **0.010** |
|  |  |  |  |  |  | Group | **0.038** |
|  | Post | 0.0 ± 1.0 | 0.0 ± 2.0 | 0.0 ± 0.0 | 0.0 ± 0.0 | Condition | **0.007** |
|  |  |  |  |  |  | Interaction | **##** |
| Vigor | Pre | 8.0 ± 5.0 | 9.0 ± 3.0 | 9.0 ± 3.0 | 9.0 ± 6.0 | Time | **<0.001** |
|  |  |  |  |  |  | Group | 0.487 |
|  | Post | 7.0 ± 4.0 | 9.0 ± 6.0 | 8.0 ± 5.0 | 9.0 ± 6.0 | Condition | 0.332 |
|  |  |  |  |  |  | Interaction | none |

Main time effect time was showed for depression [MD=0.4; 95%CI=[0.1;0.7]; F(1,32.3)=7.53; *p*=0.01], fatigue [MD=1.5: 95%CI=[1.0;2.1]; F(1,58.4)=30.4; *p*<0.001], tension [MD=0.7; 95%CI=[0.2;1.2]; F(1,39.5)=7.35; *p*=0.01] and vigor [MD=1.6; 95%CI=[0.9;2.3]; F(1,65.8)=20.5; *p*<0.001] subscales, with scores of fatigue significantly increased after the fatiguing tasks while a significant decrease was revealed for scores of depression, tension and vigor.

Main condition effect was revealed for confusion [MD=0.7; 95%CI=[0.3;1.1]; F(1,38.6)=10.8; *p*=0.002] and tension [MD=0.9; 95%CI=[0.2;1.4]; F(1,36.7)=8.09; *p*=0.007] subscales, with scores significantly higher in DT than in ST condition.

#### 3.7.3) NASA-TLX

LMM revealed that overall score from NASA-TLX questionnaire was significantly higher after DT than after ST condition [MD=6.5; 95%CI=[0.4;12.6]; F(1,24)=4.79; *p*=0.039] without main group effect [MD=6.0; 95%CI=[−5.7;17.7]; F(1,24)=1.13; *p*=0.299] or interaction effect [F(1,24)=1.03; *p*=0.320] (pwCOPD ST : 37.8±14.2; pwCOPD DT : 47.3±19.0; CTL ST : 34.8±13.7; CTL DT 38.3±17.5).

Mental demand was significantly higher after DT than after ST condition [MD=2.5; 95%CI=[0.6;4.4]; F(1,24)=7.27; *p*=0.013] without main group effect [MD=0.3; 95%CI=[−2.2;2.9]; F(1,24)=0.08; *p*=0.785] or interaction effect [F(1,24)=0.002; *p*=0.967] (pwCOPD ST : 3.1±3.5; pwCOPD DT : 5.5±5.1; CTL ST : 2.7±3.0; CTL DT 5.2±4.1).

Temporal demand was significantly higher after DT than after ST condition [MD=2.9; 95%CI=[1.1;4.8]; F(1,24)=10.63; *p*=0.003] without main group effect [MD=0.4; 95%CI=[−2.7;3.5]; F(1,24)=0.06; *p*=0.801] or interaction effect [F(1,24)=0.60; *p*=0.447] (pwCOPD ST : 4.2±3.8; pwCOPD DT : 7.8±5.3; CTL ST : 4.5±4.5; CTL DT 6.7±4.2).

No main condition, group or interaction effect was found for physical demand, performance, effort and frustration subscales.

## 4) DISCUSSION

The current study aimed at evaluating the impact of adding a working memory task during prolonged submaximal isometric contractions of knee extensors on muscle endurance and the associated modulations in neuromuscular function, cognitive control and perceived fatigue in pwCOPD compared to age- and sex-matched healthy participants. To our knowledge, this is the first study to explore muscle endurance and the associated neuromuscular adjustments in pwCOPD in dual-task context, providing novel insights into how and why fatigue develops in COPD in situations close to those which can be encountered in daily life. Contrary to our initial hypothesis, adding a low concurrent cognitive demand (i.e., 1-back task) to prolonged isolated muscle exercise did not decrease muscle endurance in both pwCOPD and healthy participants. Nevertheless, while there was no significant difference in TTF between single and dual-task conditions, pwCOPD exhibited a decrease in cognitive performance at the end of the fatiguing dual-task contrary to healthy participants who were able to maintain cognitive control despite progressive development of neuromuscular fatigability. This finding suggests an exacerbated dual-task interference under fatigue state in pwCOPD.

### 4.1) Effect of COPD on muscle endurance

As expected, regardless of the experimental conditions, pwCOPD showed reduced TTF compared to healthy participants. This result corroborates the decreased in muscle endurance in pwCOPD reported in previous studies (for review, see Evans et al. 2015). Our comprehensive assessment of fatigue and its potential contributors provide further insights on the mechanisms underlying lower muscle endurance in COPD.

In its contemporary interpretation (Enoka and Duchateau 2016), fatigue arises from the emergence and interaction of two primary forms of fatigability: (1) performance fatigability, which encompasses the peripheral and central factors underlying reduced muscle performance (e.g., calcium kinetics, voluntary activation) that can be objectively assessed, and (2) perceived fatigability, which is driven by homeostatic and psychological factors and that is often quantified using scales or questionnaires. In accordance with this model (and its adapted version in people with chronic respiratory disorders (Gruet 2018)), several factors could explain the reduced TTF in pwCOPD reported in our study.

Firstly, our results suggest that the reduction in muscle endurance in pwCOPD is, at least in part, caused by peripheral muscle mechanisms, as revealed by the early decrease in Tw_p_ amplitude (i.e., significant reduced Tw_p_ amplitude in pwCOPD compared to healthy participants at 100% time point). The importance of peripheral parameters is also indirectly confirmed by the absence of between-group differences for the kinetics of central fatigue (i.e. similar changes in VAL). While reduced central drive has been proposed as a limiting factor to physical exertion in various cardiorespiratory disorders (Marillier et al. 2022), it is of note that most evidence is derived from studies including severe patients (e.g., Marillier et al. 2018, 2022), with thus increasing likeliness of a role of chronic hypoxemia inducing cerebral impairments that are exacerbated during physical exercise. The role of central factors in limiting exercise performance is probably much more limited in moderate severity profile. For instance, our findings are in line with a previous study showing similar kinetics of central fatigue during quadriceps isometric exercise in mild-to-moderate cystic fibrosis compared to healthy controls (Gruet et al. 2016).

Our results also revealed a faster attainment of an important perceived muscle fatigue in pwCOPD in comparison with control participants. It is of note that we did not measure perceived exertion in the present study to limit the number of scores and avoid the risk of scores contamination, potentially decreasing the accuracy of each rating (Gruet et al. 2018; Lewthwaite et al. 2021). However, as effort perception is one of the main drivers of perceived muscle fatigue (Behrens et al. 2023) and because both are very closely related during motor exercise inducing progressive fatigue development (Micklewright et al. 2017), we might expect higher effort perception in pwCOPD close to task failure, as compared to healthy controls. Previous studies in healthy participants using similar exercise modalities (e.g. low-intensity sustained isometric contractions of the knee-extensors) demonstrated that task failure was related to earlier attainment of maximal perceived effort (Souron et al. 2020). Collectively, these findings suggest that higher perceived muscle fatigue / effort may have contributed to earlier disengagement from the motor task in pwCOPD, explaining, at least in part, their lower TTF.

Moreover, outcomes from the POMS questionnaire suggest more elevated levels of anger, fatigue, and tension among pwCOPD compared to healthy controls. It is of note that beyond the numerous pathophysiological factors that may contribute to the higher muscle susceptibility to fatigue in COPD (see Maltais et al. (2014) for review), lower physical activity levels (supported by Baecke questionnaire results) and compromised sleep quality (supported by PSQI questionnaires results) may have also played a role through their negative influence on various modulating factors of both perceived (e.g. wakefulness, arousal) and performance fatigability (e.g. force capacity) (Enoka and Duchateau 2016; Gruet 2018).

Hence, it appears that the reduced TTF observed in pwCOPD is attributed to a combination of psychological and physiological factors.

### 4.2) Effect of CMDT on muscle endurance

Contrary to our expectations, we did not identify a difference in TTF between ST and DT conditions, for both pwCOPD and healthy individuals. This finding is in contrast to the results reported by Chatain et al. (2019) in young healthy adults, showing a reduced muscle endurance when participants performed a 1-back task concomitantly to a motor task relatively close to that used in the present study (i.e., intermittent submaximal contractions of knee extensors performed at 20% MVF until exhaustion). Nevertheless, differences in protocols characteristics could explain the discrepancies in the results.

The duration of muscle contractions performed during motor task in Chatain et al. (2019) was longer than in the present study (i.e., sets of contractions of 170s vs. 70s in the present study), for identical neuromuscular evaluation timings (i.e., 40s) during which no cognitive task was performed. Therefore, participants were exposed to the 1-back task 75% of the exercise time in Chatain et al. (2019), compared to 55% in the present study. This reduced time exposure to the cognitive task may have impede the development of mental fatigue, which has been suggested as one of the main contributors to the decline in muscle endurance in CMDT (Chatain et al. 2019). This assumption is reinforced by the scores obtained from the mental demand subscale of the NASA-TLX questionnaire which, although significantly different between ST and DT, remain relatively low compared to other studies (Shortz and Mehta 2017; Chatain et al. 2019). It is of note that participants started the DT condition with a level of cognitive performance lower that the maximum achievable (i.e., D’_0%_=2.87 vs. D’_max_=3.47, i.e., 83% of maximum cognitive performance). Thus, from the beginning of the task, participants were not able to perform the cognitive task maximally, probably in order to focus on the motor task, which could mask the detrimental effects of the cognitive task on muscle endurance.

While in one hand the cognitive load was not sufficient to have detrimental effect on endurance time, in the other hand the cognitive task was enough difficult to reveal the inability of pwCOPD to maintain cognitive performance through the endurance task as they probably allocate more cerebral resources to maintain motor performance.

### 4.3) Effect of CMDT and COPD on muscle endurance

Despite the absence of an (group x condition) interaction effect on muscle endurance, we observed an average TTF reduction in DT condition of −11.2% among pwCOPD while it showed a slight increase of 0.9% in healthy participants.

Beyond the aforementioned questionable engagement of the participants in the cognitive task, the absence of a statistically significant interaction effect could be attributed to an important interindividual variability in endurance times or also explained by the significant decrease of cognitive performance observed at the end of the fatiguing task in pwCOPD compared to controls. Indeed, the D’ values decreased significantly only in pwCOPD from 100% time point. It is of note that this decrease in cognitive performance occurred despite unchanged reaction time and non-answer, suggesting that they do not deliberately give up on the cognitive task (which may have resulted in a decrease in reaction time and/or an increase in number of non-answers) but probably allocate more resources to the maintenance of the motor task. Therefore, it can be hypothesized that if pwCOPD would have maintained a constant level of cognitive performance, this might have compromised the motor task, leading to decreased muscle endurance in DT condition. In any case, the reduced cognitive performance of pwCOPD at the end of prolonged CMDT reflects a higher level of interference, corroborating the data from previous work in brief unfatiguing tasks (Morlino et al. 2017; Heraud et al. 2018; Ozsoy et al. 2021) and extending this observation in a fatiguing context.

Nevertheless, while cognitive performance was reduced in pwCOPD in the presence of muscle fatigue, our results showed that without fatigue (i.e., during the first steps of the fatiguing tasks) pwCOPD exhibited similar DT performance compared to healthy participants (i.e., similar performance in the 1-back task while maintaining similar submaximal relative force), contrasting with previous studies (Morlino et al. 2017; Heraud et al. 2018; Ozsoy et al. 2021). This result can be explained by the fact that the motor task used in the current study is based on isolated muscle contractions while previous studies used whole-body tasks (e.g., walking task, TUG test). This finding suggests that in the absence of fatigue, the presence of motor dual-task interference is directly related to the nature of the motor and/or cognitive task. Considering this point and our results showing that fatigue can exacerbate dual-task interference for pwCOPD, we can hypothesize that the effects of fatigue could be even greater when DT involves a whole-body motor task.

### 4.4) Clinical implications

Both cognitive function and physical fitness are impaired in pwCOPD (Dodd et al. 2010; Maltais et al. 2014). Exercise training are already included in the package of care of pwCOPD with accumulating evidence regarding positive effect on both neuromuscular function and cardiorespiratory fitness (Spruit et al. 2016). Meanwhile, interest for cognitive training is more recent, with very few trials (Incalzi et al. 2008; van Beers et al. 2021) showing only limited benefits when performed in isolation (e.g., unchanged psychological wellbeing and healthy lifestyle goals following 12-week of working memory training (van Beers et al. 2021)). It is also unclear whether training motor and cognitive functions in isolation may translate in improvements in cognitivo-motor processing during dual tasking, and ultimately improve ability to better perform daily living tasks. Moreover, perceived lack of time or competing priorities are important barrier to structured physical activity among older adults (Kosteli et al. 2016), including pwCOPD and notably those in full-time employment (Kosteli et al. 2017). In this context, CMDT training may (1) serve as a time-effective strategy to improve both motor and cognitive functions and (2) present the advantage to tap into cerebral resources that are not fully engaged when the two tasks are performed independently, as suggested by the exacerbated dual-task interference under fatigue state in pwCOPD.

Our results also provide evidence on the feasibility to add a cognitive challenge during prolonged fatiguing physical exercise in pwCOPD, which was important to confirm as producing fatigue is a prerequisite for optimal results from an exercise training program (e.g., Burtin et al. 2012). Implementing a dual-task paradigm may be particularly interesting for resistance training, which is an important component of exercise training in COPD (Spruit et al. 2002; Gloeckl et al. 2013), aiming to increase muscle strength and endurance while keeping ventilatory requirements tolerable. Our results are particularly relevant in this context as the working memory task was added to a single-joint exercise of the most targeted muscle group (i.e., quadriceps) in resistance training sessions in COPD. However, it is of note that implementing cognitive task to whole body exercise (e.g., aerobic cycling exercise) may be difficult as increased dyspnea perception may compromise, at some points during the session, the ability to focus on the concomitant cognitive task.

Finally, our findings suggest that adding cognitive challenge does not compromise the motor task in terms of muscle fatigability and task duration which are important components to elicit muscle adaptations while adding a concurrent cognitive challenge. Of course, the exact difficulty and duration of both cognitive and motor tasks remain to be adjusted for real exercise training sessions.

### 4.5) Conclusion

In conclusion, our study revealed that COPD did not induce a larger decrease in muscle endurance in presence of concomitant cognitive task compared to a control condition. Although neuromuscular fatigability was not affected by the addition of cognitive demand, pwCOPD showed reduced cognitive performance at the end of the fatiguing dual-task, reflecting an increase of interference in a fatigue situation for these patients. These findings highlight the importance of considering the integration of fatiguing CMDT in rehabilitation programs to fully solicit the available cerebral resources and ultimately potentially increase the expected benefits, especially in cerebral function. Nevertheless, more research is needed to fully understand COPD effect on fatigability during prolonged dual-task, the underlying mechanisms involved and the interest of including these procedures in rehabilitation programs.

## Author contributions

**Cyril Chatain**: Conceptualization; Data curation; Formal analysis; Investigation; Methodology; Visualization; Writing – original draft; Writing – review & editing. **Jean-Marc Vallier**: Conceptualization; Funding acquisition; Investigation; Supervision; Writing – review & editing. **Nicolas Paleiron**: Conceptualization; Investigation; Resources; Writing – review & editing. **Fanny Cucchietti**: Data curation; Validation; Project administration; Funding acquisition; Writing – review & editing. **Sofiane Ramdani**: Conceptualization; Funding acquisition; Writing – review & editing. **Mathieu Gruet**: Conceptualization; Funding acquisition; Methodology; Supervision; Writing – original draft; Writing – review & editing.

## Acknowledgments

This study was supported by a doctoral research grant (recipient Cyril Chatain) from the French Ministère de l’Enseignement Supérieur, de la Recherche et de l’Innovation (MESRI) and by funds from the Centre Hospitalier Intercommunal Toulon – La Seyne-sur-Mer (CHITS). The authors declare to have no financial, personal or other conflicts of interest.

## Data availability statement

Data can be obtained from the first author (Cyril Chatain) upon reasonable request.

